# Brain volumes and their relationship with cerebral microbleeds and cognition in middle-aged adults with type 1 diabetes

**DOI:** 10.64898/2026.08.04.26359672

**Authors:** Iiris Kyläheiko, Linda Kuusela, Tor-björn Claesson, Aleksi Tarkkonen, Juha Martola, Teemu I. Paajanen, Jussi Virkkala, Per-Henrik Groop, Lena M. Thorn, Turgut Tatlisumak, Jukka Putaala, Daniel Gordin, Hanna Jokinen, the FinnDiane Study Group

## Abstract

**Objective:** Type 1 diabetes is related to an increased risk of structural brain alterations, cerebral microbleeds (CMBs), and cognitive deficits. We explored brain volumes and their direct and combined associations with CMBs on cognitive performance in middle-aged individuals with type 1 diabetes.

**Research Design and Methods:** Adults with type 1 diabetes (n=163; mean age 46±8 years; diabetes duration 31±10 years; 53% women) and 48 matched controls underwent brain MRI and clinical and neuropsychological evaluations. Volumetric MRI measures adjusted to intracranial volume included total brain volume (TBV), white matter volume (WMV), and total volumes of cortex, thalamus, hippocampus, nucleus accumbens, and choroid plexus.

**Results:** Individuals with type 1 diabetes had smaller TBV, WMV, and volumes of cortex, thalamus, and nucleus accumbens, and larger choroid plexus compared to controls (Cohen’s *d*=0.39–0.54). Those with type 1 diabetes and ≥3 CMBs had smaller TBV, WMV, and volumes of cortex, thalamus, and nucleus accumbens, compared to those with 0–2 CMBs (Cohen’s *d*=0.54–0.92). We found no direct associations between brain volumes and processing speed or executive functions. However, TBV, WMV, nucleus accumbens, and choroid plexus volumes had significant negative synergistic interactions with CMBs on processing speed and executive functions (standardized betas: −0.61 to −0.51 and 0.54 to 0.75, p^FDR^=0.006–0.048).

**Conclusions:** Smaller global and regional brain volumes and larger choroid plexus volumes were found in middle-aged individuals with type 1 diabetes compared to healthy controls. Together with CMB burden, structural brain volumetric alterations were associated with accelerated cognitive deficits.

**Twitter summary:** In middle-aged adults with #Type1Diabetes, global and regional brain volumes together with cerebral microbleeds are associated with accelerated cognitive deficits. #Cognition #BrainMRI

**Article highlights:**

- Why did we undertake this study? The interplay between cerebral small vessel disease, brain volumetric alterations, and cognition in type 1 diabetes remains unclear.
- What is the specific question(s) we wanted to answer? Does type 1 diabetes involve brain volume loss in middle age, is this loss related to cerebral microbleed (CMB) burden or cognitive deficits, and does CMB burden moderate the association between brain volume and cognition?
- What did we find? Several brain volumetric alterations were found, which were not directly related to cognitive changes. However, we observed negative synergistic interactions between brain volumetric changes and CMBs on cognitive outcomes.
- What are the implications of our findings? Microvascular pathology and structural brain volumetric alterations jointly contribute to cognitive impairment in type 1 diabetes.

## 1 INTRODUCTION

Type 1 diabetes has adverse effects on the development of brain and cognition in childhood and youth ^1^. Later on, in young adulthood as well as in midlife and older age, type 1 diabetes poses a risk for cerebral volumetric ^2,3^, microstructural ^4^, microvascular ^5,6^, and functional ^7^ changes, as well as for cognitive deficits ^8^.

In middle-aged adults with type 1 diabetes, the most consistent evidence for brain volumetric alterations concerns thalamic atrophy ^2^, though reductions in global and other regional brain volumes ^3,9–12^ have also been reported. However, contradictory findings exist ^9,11,13,14^. Risk factors for volume loss include older age ^11,13,15^, cardiovascular risk factors ^3,11^, hyperglycemia ^3,16^, diabetes duration ^11^, and concomitant microvascular disease ^3,13^, including white matter hyperintensities (WMH) ^12^. Smaller global ^3,9^ and regional ^11,12^ brain volumes have in turn been linked to poorer cognitive performance, particularly in processing speed, attention, and executive functions, although evidence remains limited.

Cerebral small vessel disease (cSVD) may represent an important mechanism linking structural brain changes to cognitive dysfunction in type 1 diabetes. In our cohort of middle-aged adults with type 1 diabetes, cerebral microbleeds (CMBs), unlike mild WMHs, were associated with early cognitive deficits ^17^. Furthermore, individuals with any cSVD marker exhibited smaller volumes of total white matter (WMV), deep gray matter (GMV), cortex, thalamus, and basal ganglia compared with those without cSVD. Nevertheless, the specific contribution of CMB burden ^5,18^ to brain volume loss and its potential role in modifying the relationship between structural brain alterations and cognition remains unknown. To date, only one study has examined the interplay between volumetric, microvascular, and cognitive changes in type 1 diabetes and found no moderating effect of WMHs on the association between GMV and processing speed ^12^. Another potentially relevant but largely unexplored marker is choroid plexus volume. Emerging evidence links enlargement of the choroid plexus to glymphatic dysfunction ^19^, cSVD ^20^, cSVD-related cognitive impairment ^21^, neuroinflammation ^22^, and mild cognitive impairment ^23^. Despite the established presence of cSVD in type 1 diabetes, choroid plexus has not yet been investigated in this population.

The present study examined structural brain volumetric alterations and their associations with cognitive performance in middle-aged adults with type 1 diabetes. We hypothesized that individuals with type 1 diabetes would exhibit reduced global and regional brain volumes compared with healthy controls and that these volumetric changes would be associated with poorer performance in processing speed and executive functioning. We further explored whether greater CMB burden is associated with more pronounced volumetric abnormalities and whether CMBs moderate the associations between brain volumes and cognition.

## 2 RESEARCH DESIGN AND METHODS

### 2.1 Participants and study protocol

This MRI sub-study on early markers for cerebrovascular disease ^18^ is a part of the longitudinal, nationwide Finnish Diabetic Nephropathy (FinnDiane) Study (all study centers described in Supplemental Table 1) ^5,17^. Initial inclusion criteria at enrolment in 2011–2017 were age 18–50 years, onset of diabetes before age 40, and no kidney replacement therapy, history of overt neurovascular disease, assessed with the Questionnaire for Verifying Stroke-Free Status ^24^, or contraindications for brain MRI ^18^. The current study focuses on cross-sectional data collected at follow-up carried out during 2019–2024 (Supplemental Figure 1). Exclusion criteria for the current analyses included neurological diseases that could affect the brain and cognition. The ethics committee of Helsinki University Hospital approved the study, and it was conducted according to the Declaration of Helsinki. All participants signed an informed consent.

### 2.2 Clinical examinations

Medical history, medications, office blood pressure, waist circumference, and body mass index were recorded (Table 1) at the FinnDiane Research Unit, Helsinki University Hospital. Blood samples were analyzed for glycated hemoglobin (HbA_1c_), lipids, and creatinine. Estimated glomerular filtration rate (eGFR) was determined with the Chronic Kidney Disease Epidemiology Collaboration 2009 equation. Albuminuria was defined as moderately or severely increased albuminuria based on two out of three urine collections for participants with type 1 diabetes and on a single 24h urine collection for controls. Retinal photocoagulation was used as an indicator of severe diabetic retinopathy. Estimated glucose disposal rate (eGDR), a marker of insulin resistance, was calculated with the following formula: eGDR = 24.4 - (12.97 × WHR) - (3.39 × AHT) - (0.60 × HbA_1c_); WHR = Waist-hip-ratio; AHT = yes/no (yes = SBP ≥ 140 or DBP ≥ 90 or antihypertensive treatment) ^25^. Demographic and lifestyle information was collected via questionnaires ^18^.

**Table 1.** Demographic, clinical, and brain imaging characteristics of controls and individuals with.

|  | Controls (n=48) | Individuals with type 1 diabetes (n=163) | p-value |
| --- | --- | --- | --- |
| Age, years | 46.3 (10.1) | 46.4 (7.7) | 0.903 |
| Female sex, n (%) | 25 (52.1) | 87 (53.4) | 1.000 |
| Education, years | 17.2 (4.0) | 16.8 (3.3) | 0.474 |
| Age at diabetes onset, years | NA | 15.4 (9.3) | NA |
| Diabetes duration, years | NA | 31.4 (10.4) | NA |
| BMI, kg/m<sup>2</sup> | 26.0 (4.8) | 28.0 (4.9) | **0.003** |
| Waist-to-height ratio | 0.52 (0.07) | 0.55 (0.08) | **0.003** |
| Systolic blood pressure, mmHg | 129 (16) | 131 (17) | 0.247 |
| Diastolic blood pressure, mmHg | 83 (10) | 82 (10) | 0.299 |
| Antihypertensive medication, n (%) | 6 (12.5) | 73 (44.8) | **<0.001** |
| HbA<sub>1c</sub>, mmol/mol | 34 (3) | 59 (10) | **<0.001** |
| HbA<sub>1c</sub>, % | 5.3 (0.3) | 7.6 (0.9) | **<0.001** |
| Total cholesterol, mmol/L | 4.9 (0.8) | 4.1 (0.9) | **<0.001** |
| LDL cholesterol, mmol/L | 3.0 (0.7) | 2.2 (0.8) | **<0.001** |
| HDL cholesterol, mmol/L | 1.4 (0.4) | 1.5 (0.4) | 0.312 |
| Triglycerides, mmol/L | 1.1 (0.6) | 1.1 (0.6) | 0.622 |
| Lipid-lowering medication, n (%) | 1 (2.1) | 98 (60.5) | **<0.001** |
| Coronary heart disease, n (%) | 0 (0.0) | 6 (3.7) | 0.341 |
| Current smoker, n (%) | 7 (14.9) | 9 (5.6) | 0.071 |
| Albuminuria, n (%) | 0 (0) | 34 (20.9) | **0.001** |
| eGFR, ml/min/1.73m<sup>2</sup> | 100 (14) | 102 (16) | 0.262 |
| Diabetic retinopathy, n (%) | 0 (0.0) | 49 (30.1) | **<0.001** |
| CMB burden (≥3) | 1 (2.1) | 25 (15.3) | **0.027** |
| WMH volume, cm<sup>3</sup> | 0.65 (0.46) | 1.35 (2.03) | **0.026** |
| Lacunar infarcts, n (%) | 0 (0.0) | 3 (1.8) | 1.000 |

Data are presented as mean (SD), if not mentioned otherwise. BMI, body mass index; CMB, cerebral microbleed; eGFR, estimated glomerular filtration rate; HbA<sub>1c</sub>, glycated hemoglobin; HDL, high-density lipoprotein; LDL, low-density lipoprotein; WMH, white matter hyperintensity.

### 2.3 Brain MRI

Brain MRI was performed at the Helsinki University Hospital’s Medical Imaging Center with 3T Philips Ingenia scanner (Best, The Netherlands) with a 32-channel head coil ^5^. Sequences included T1, T2, FLAIR, susceptibility-weighted imaging, T2*, diffusion-weighted imaging, T1 Magnetization-Prepared Rapid Gradient Echo, and MR time-of-flight. Senior neuroradiologist (JM) evaluated visually CMBs, WMHs (Fazekas scale), infarcts, and lacunes according to standard criteria ^26,27^, blinded to clinical and neuropsychological data. Individuals with type 1 diabetes were divided into two groups based on their CMB number (0–2 vs ≥3) ^17^.

The analyses included both global and regional MRI volume markers previously shown to differentiate individuals with type 1 diabetes and/or cSVD and controls, including total brain volume (TBV) ^3^, WMV ^3,9^, and volumes of cortex ^10^, thalami ^2^, hippocampi ^12,26^, nucleus accumbens ^11^, and lateral choroid plexuses ^20,21^. These volumetric markers as well as WMHs for descriptive purposes were segmented from 3D FLAIR images using the Lesion Prediction Algorithm ^28^, integrated into the Lesion Segmentation Tool for SPM (version 3.0.0; www.statistical-modelling.de/lst.html). To correct for individual differences in brain size, all MRI volumetric markers were normalized by dividing them by the total intracranial volume ^29^, which was obtained through segmentation of the T1 and T2 3D TSE sequences using Freesurfer (version 7.1.1) ^30,31^. Any required adjustments to the segmentations were made manually by a proficient physicist (LK). As Freesurfer does not reliably perform segmentation of the choroid plexus, we performed its segmentations using ASCHOPLEX ^32^, a machine learning model developed for choroid plexus segmentation. As visually inspected, all segmentations were of acceptable quality.

### 2.4 Neuropsychological assessment

The neuropsychological protocol has been described in earlier publications ^17^ and in Supplemental table 2. In short, trained research psychologists provided neuropsychological assessments including both established neuropsychological tests and computerized cognitive tests. Measures of processing speed and executive functions for the current analyses were selected based on earlier literature ^8^, and our earlier research findings in the same cohort showing them to be most sensitive to cognitive changes ^17^. Processing speed was evaluated with WAIS-IV Coding, Stroop Color-Naming (Stroop- II), and computerized Flexible Attention Test (FAT) ^33^ subtask of Reaction Time (FAT-RT).

Executive function subdomain of inhibitory control was covered with Stroop Color-Incongruent (Stroop-III), cognitive flexibility with FAT Number Month Backward (FAT-NMB) and working memory with FAT Visuospatial Memory Span Backward (FAT-MB) ^33^. Hypoglycemic events were to be avoided 24h prior to the assessment. Glucose level ≥3 mmol/L was required before the examination and was measured with capillary test or glucose monitoring sensor, if available.

### 2.6 Statistical analysis

Descriptive analyses and group comparisons were performed with Spearman correlation and Student t-test after checking the assumptions, where appropriate. Data are reported as mean (SD), if not indicated otherwise. Missing clinical or cognitive data, which was due to the participant’s failure to complete the task, or technical or other reasons, were not replaced.

We performed univariate linear regression models to assess the associations between the key demographic, diabetes-related, and vascular risk factors and MRI volumetric markers in participants with type 1 diabetes. The selected clinical variables for these models included age, sex, diabetes duration, HbA_1c_, waist/height ratio, systolic and diastolic blood pressure, antihypertensive medication, total cholesterol, high- and low-density lipoproteins, triglycerides, lipid-lowering medication, eGDR, smoking, diabetic retinopathy, and eGFR. We further assessed the associations between clinical markers and brain volumes with multivariate linear regression models, where the main clinical variables were substituted with eGDR to reduce the number of independent variables. As sensitivity analyses, we added diabetes-related complications, i.e., albuminuria and diabetic retinopathy, into the multivariate model. Next, we examined how MRI volumetric markers are associated with cognition in age- and sex-adjusted models. Subsequently, Student t-tests were performed to test whether participants with type 1 diabetes and ≥3 CMBs ^17^, the most prominent cSVD marker in our cohort ^5,18^, differ from those with 0–2 CMBs in brain volumes. Finally, moderating effects of CMBs on associations between volumetric brain measures and cognition were assessed by including the interaction term (MRI volume marker × CMB) into the age- and sex-adjusted multivariate models with cognition as dependent variable. As sensitivity analyses, interaction models were run without visually estimated influential data points.

To facilitate comparability of regression coefficients across predictors, all continuous variables were z-standardized prior to inclusion in the models. Additionally, the z-scores for FAT-RT, FAT-NMB, Stroop-II, and Stroop-III were reversed so that higher scores consistently indicated better performance across all cognitive tests. Logarithmic transformations were performed for dependent variables, where appropriate. In the multivariable linear regression models, no systematic evidence for violation of homoscedasticity was detected. A quarter of the models showed a slight violation of the residual normality assumption due to outliers, but Cook’s distance for influential data points remained consistently below one. Statistical significance was set at p<0.05. Effect sizes were estimated using Cohen’s *d.* To control multiplicity, false discovery rate (FDR) correction was applied for interaction models using the Benjamini-Hochberg procedure. All analyses were conducted in R (version 4.5.1).

## 3 RESULTS

### 3.1 Participant characteristics

The analyses were performed on 163 participants with type 1 diabetes and 48 healthy controls matched by age, sex, and education (Supplemental Figure 1), after excluding 7 individuals with type 1 diabetes (traumatic brain injury, n=3; multiple sclerosis, n=2; cerebral infarct, n=2) and one control (dementia, n=1). Descriptive data of individuals with type 1 diabetes and controls are presented in Table 1. Among participants with type 1 diabetes, 88.5% (n=138) had glucose concentrations prior to the neuropsychological evaluation between 4.0–13.9 mmol/L, 9.0% (n=14) had ≥14 mmol/L, and 2.6% (n=4) had 3.0–3.9 mmol/L. The glucose concentration did not correlate with performance in any of the cognitive tasks (p>0.05). Median time between brain MRI and neuropsychological evaluation was 14.0 days (IQR 6.0–28.5). Number of missing values ranged in controls in clinical data (Table 1) between 1–2, and in individuals with type 1 diabetes in clinical and cognitive data and between 1–3, except for FAT-MB (n=17). Participants with type 1 diabetes and available FAT-MB values did not differ from those without in age, sex, or years of education, but had lower HbA_1c_ on average, compared to those without (59 vs 64 mmol/mol, respectively, p<0.05).

### 3.2 Brain MRI volume comparisons between individuals with type 1 diabetes and controls

Compared to healthy controls, adults with type 1 diabetes had smaller TBV, WMV, and volumes of cortex, thalamus, and nucleus accumbens, and larger choroid plexus (Table 2). No differences were found for hippocampus. Effect sizes for the significant group differences were small to moderate or moderate in magnitude.

**Table 2.** Brain MRI volumes in controls and individuals with type 1 diabetes, and in cerebral microbleed (CMB) groups among participants with type 1 diabetes.

|  | All participants (n=211) |  |  |  | Individuals with type 1 diabetes (n=163) |  |  |  |
| --- | --- | --- | --- | --- | --- | --- | --- | --- |
|  | Controls<br>(n=48) | Individuals<br>with type 1<br>diabetes<br>(n=163) | Cohen's *d* | p-value | 0-2 CMBs<br>(n=138) | ≥3 CMBs<br>(n=25) | Cohen's *d* | p-value |
| TBV (cm<sup>3</sup>) | 1178.40<br>(102.90) | 1144.55<br>(113.14) |  |  | 1143.49<br>(113.15) | 1150.41<br>(115.26) |  |  |
| % of ICV | 72.09<br>(2.54) | 70.68<br>(2.65) | 0.54 | **0.001** | 71.03<br>(2.58) | 68.81<br>(2.27) | 0.87 | **<0.001** |
| WMV (cm<sup>3</sup>) | 477.00<br>(50.89) | 460.68<br>(59.12) |  |  | 460.37<br>(59.25) | 462.38<br>(59.59) |  |  |
| % of ICV | 29.17<br>(1.74) | 28.39<br>(1.77) | 0.44 | **0.008** | 28.54<br>(1.78) | 27.59<br>(1.50) | 0.54 | **0.014** |
| Cortex<br>volume (cm<sup>3</sup>) | 496.47<br>(45.24) | 482.40<br>(44.87) |  |  | 482.19<br>(45.09) | 483.62<br>(44.53) |  |  |
| % of ICV | 30.37<br>(1.26) | 29.82<br>(1.45) | 0.39 | **0.013** | 29.98<br>(1.46) | 28.95<br>(1.01) | 0.73 | **<0.001** |
| Thalamus<br>volume (cm<sup>3</sup>) | 16.31<br>(1.70) | 15.58<br>(1.62) |  |  | 15.64<br>(1.61) | 15.25<br>(1.62) |  |  |
| % of ICV | 1.00<br>(0.07) | 0.96<br>(0.07) | 0.50 | **0.005** | 0.97<br>(0.07) | 0.91<br>(0.06) | 0.92 | **<0.001** |
| Hippocampus<br>volume (cm<sup>3</sup>) | 8.63<br>(0.91) | 8.54<br>(0.78) |  |  | 8.53<br>(0.78) | 8.59<br>(0.79) |  |  |
| % of ICV | 0.53<br>(0.05) | 0.53<br>(0.04) | 0.01 | 0.970 | 0.53<br>(0.04) | 0.52<br>(0.04) | 0.42 | 0.057 |
| Nucleus<br>accumbens<br>volume (cm<sup>3</sup>) | 0.99<br>(0.19) | 0.92<br>(0.17) |  |  | 0.93<br>(0.18) | 0.87<br>(0.16) |  |  |
| % of ICV | 0.06<br>(0.01) | 0.06<br>(0.01) | 0.36 | **0.029** | 0.06<br>(0.01) | 0.05<br>(0.01) | 0.57 | **0.010** |
| Choroid<br>plexus<br>volume (cm<sup>3</sup>) | 3.15<br>(0.78) | 3.44<br>(0.74) |  |  | 3.39<br>(0.74) | 3.69<br>(0.69) |  |  |
| % of ICV | 0.19<br>(0.05) | 0.21<br>(0.05) | -0.44 | **0.008** | 0.21<br>(0.05) | 0.22<br>(0.05) | -0.24 | 0.226 |

Data are presented as mean (SD) and represent unnormalized and normalized values. Group differences were analyzed with Student t-test for brain volumetric measures normalized to intracranial volume. TBV, total brain volume; WMV, white matter volume.

### 3.3 Associations between clinical markers and brain volumes in type 1 diabetes

Results from univariate associations between selected clinical and demographic variables and global and regional brain volumes in people with type 1 diabetes are presented in Figure 1. Age, female sex, antihypertensive medication, diabetes duration, and eGDR were most often associated with the brain volumes.

**Figure 1.**
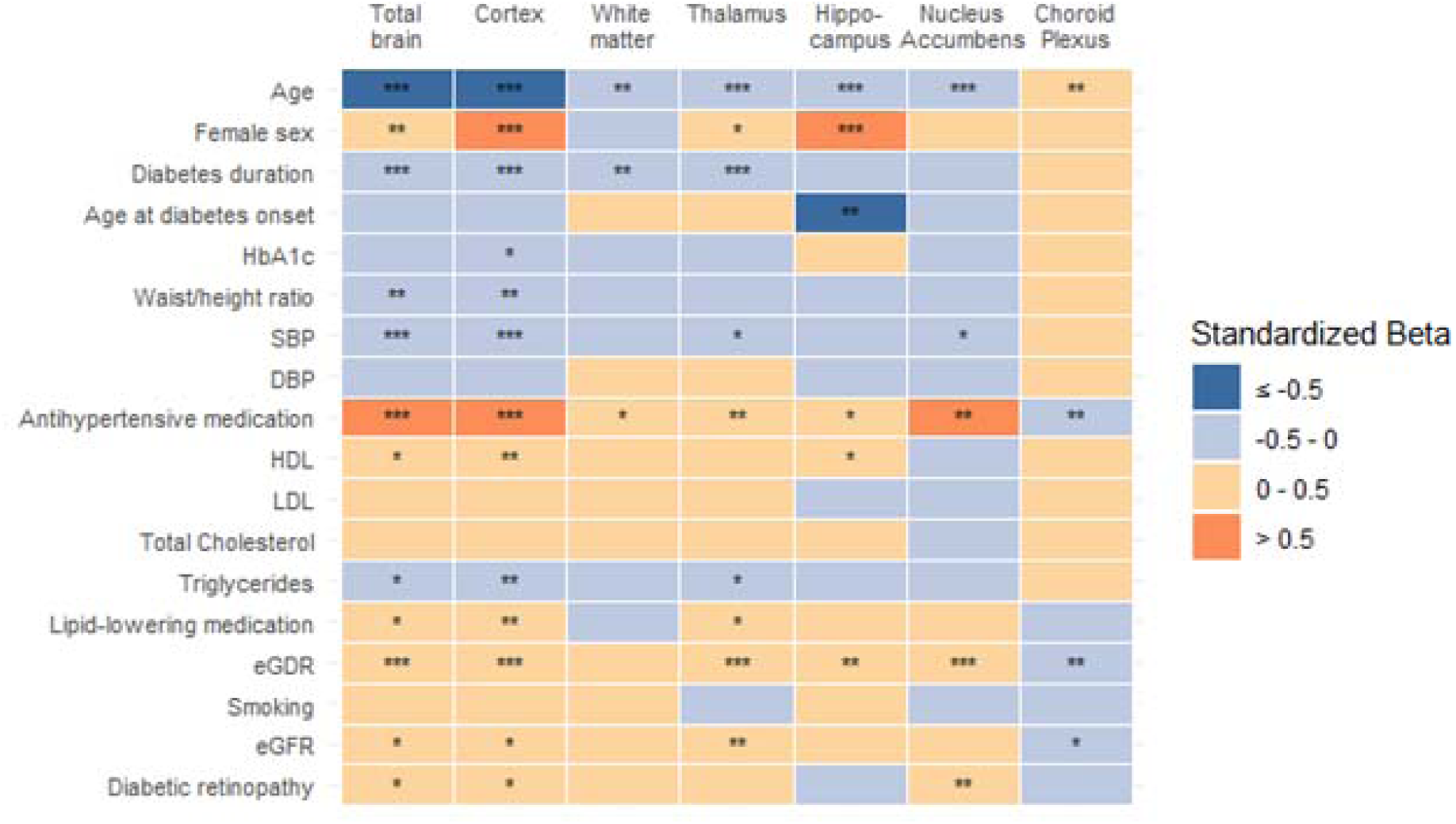
Univariate linear regression associations between normalized brain MRI volumes and selected demographic and clinical factors in individuals with type 1 diabetes. Heatmap of standardized β coefficients from univariate linear regression models with normalized brain volume measures as dependent variables and clinical and demographic variables as independent variables. Each cell represents the standardized β coefficient, categorized into four groups (≤ −0.5, −0.5 to 0 0.5, > 0.5), with blue shades indicating negative and orange shades positive associations. Significance levels are indicated by asterisks (* p < 0.05, ** p < 0.01, *** p < 0.001). For sex, β reflects female vs. male. DBP, diastolic blood pressure; eGDR, estimated glucose disposal rate; eGFR, estimated glomerular filtration rate; HDL, high-density lipoprotein; LDL, low-density lipoprotein; SBP, systolic blood pressure.

In the multivariate models, age was independently associated with all brain volumetric markers except for choroid plexus, i.e., with TBV and white matter, cortex, thalamus, hippocampus, and nucleus accumbens volumes (standardized β from −0.44 to −0.25, p≤0.005; Supplemental Table 3). Female sex was associated with larger cortex and hippocampus (standardized β 0.28–0.66, p≤0.005), and eGDR with TBV (standardized β=0.19, p=0.019), cortex (standardized β=0.30, p<0.001), and choroid plexus volumes (standardized β=-0.22, p=0.021). In sensitivity analyses adjusting additionally for diabetic complications, diabetic retinopathy was associated with nucleus accumbens (standardized β=0.33, p=0.048). The results remained the same, except for the association between sex and cortex not reaching significance (Supplemental Table 4).

### 3.4 Associations between brain volumes and cognition in individuals with type 1 diabetes

Among participants with type 1 diabetes, no direct associations were found between volumetric MRI markers and cognition (Supplemental Table 5).

### 3.5 Associations between brain volumes and CMBs in individuals with type 1 diabetes

Individuals with type 1 diabetes and ≥3 CMBs had smaller TBV, WMV, and volumes of cortex, thalamus, and nucleus accumbens, compared with those with 0–2 CMBs (Table 2). Effect sizes for the significant group differences were moderate or large in magnitude. No differences were found between hippocampus or choroid plexus volumes among the CMB groups.

### 3.6 Interactions of brain volumes and CMBs on cognition in individuals with type 1 diabetes

In the interaction models including brain volumes and CMBs as interaction terms (brain volume × CMB group), as shown in Figure 2 and Supplemental Table 6, CMBs moderated the associations between TBV and processing speed (FAT-RT, standardized β=0.54) and executive functions (Stroop-III and FAT-NMB, standardized β=0.48 and 0.75, respectively), between WMV and processing

**Figure 2.**
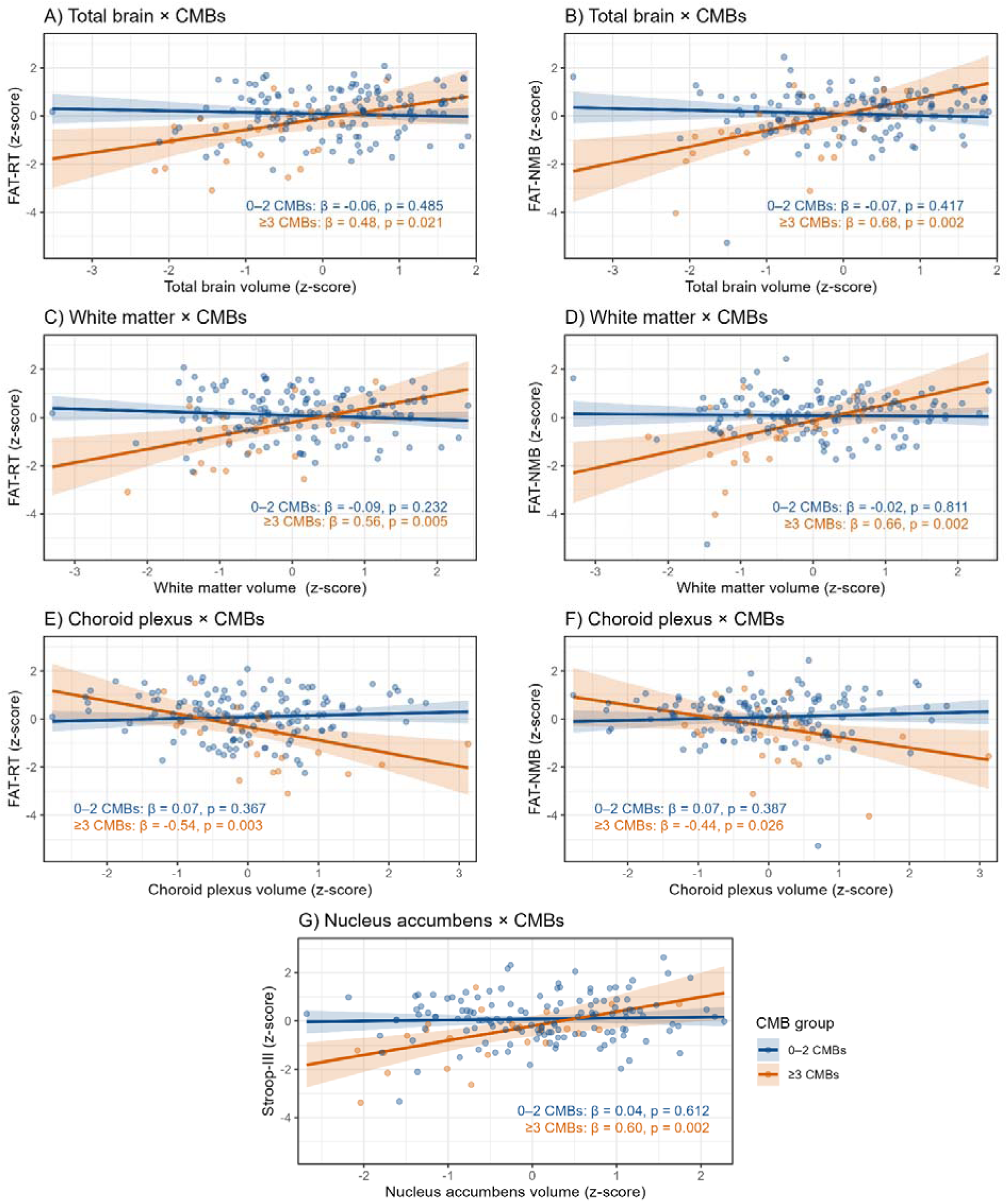
Interaction models with cognitive test scores as dependent variables, including an interaction term brain volume marker × cerebral microbleeds (CMBs), adjusted for age and sex in individuals with type 1 diabetes (n=163). Brain volumes (A–B: total brain volume, C–D: white matter volume, E–F: choroid plexus volume, and G: nucleus accumbens volume) and cognitive outcomes are presented as standardized z-scores. Z-scores for FAT-RT, FAT-NMB, Stroop-II, and Stroop-III are reversed so that higher scores consistently indicate better performance across all cognitive tests. Solid lines represent model-based predicted associations between brain volume and cognition for each CMB group (0–2 vs ≥3 CMBs), derived from interaction models. Shaded areas indicate 95% confidence intervals. Text annotations display simple slopes (standardized β coefficients) and corresponding p-values for each CMB group. All interactions were significant after false discovery rate correction. FAT, Flexible Attention Test; FAT-NMB, FAT Numbers and Months Backward; FAT-RT, FAT Reaction Time; Stroop-III, Stroop Color-Incongruent.

speed (FAT-RT, standardized β=0.65) and executive functions (Stroop-III and FAT-NMB, standardized β=0.47 and 0.68, respectively), between cortex and executive functions (FAT-NMB, standardized β=0.68), between nucleus accumbens and executive functions (Stroop-III, standardized β=0.56) and between choroid plexus and processing speed (Coding and FAT-RT, standardized β=−0.46 and −0.61, respectively) and executive functions (FAT-NMB, standardized β=−0.51). CMB group did not moderate associations between other brain volumes and cognitive performance. After FDR correction, interactions of TBV and CMBs for FAT-RT and FAT-NMB (p^FDR^=0.039 and p=0.006, respectively), of WMV and CMBs for FAT-RT and FAT-NMB (p^FDR^=0.009 for both), nucleus accumbens and CMBs for Stroop-III (p^FDR^=0.048), and of choroid plexus and CMBs for FAT-RT and FAT-NMB (p^FDR^=0.012 and p=0.048, respectively) remained significant. In all these analyses, higher CMB potentiated the effect of structural volumes on cognitive impairment. Without influential data points, the results surviving FDR correction changed only regarding the interaction of CMBs and choroid plexus volume on FAT-NMB (p=0.055).

## 4 CONCLUSIONS

This study shows that individuals with type 1 diabetes have significant brain volume loss compared to healthy controls in middle age, and that this volume loss is associated with CMB burden. We also show the enlargement of choroid plexus, which has been proposed as a new possible MRI marker of glymphatic dysfunction ^19^. Interestingly, although no direct relationships were observed between brain volumes and cognitive functions, volume loss in conjunction with CMBs demonstrated negative synergistic effects on processing speed and executive functions. Individuals exhibiting a higher number of CMBs showed more pronounced associations between total brain, white matter, and nucleus accumbens atrophy, choroid plexus enlargement, and diminished cognitive performance.

To our knowledge, this is the first study to show enlarged choroid plexus in individuals with type 1 diabetes compared with healthy controls. This finding aligns with those in cSVD ^20^ and type 2 diabetes ^34^. Choroid plexus is a highly vascularized structure that produces cerebrospinal fluid and plays a key role in waste clearance and the blood-CSF barrier ^22^. Its enlargement has been suggested to reflect glymphatic dysfunction, potentially contributing to cSVD ^19^ and cognitive dysfunction ^35^. However, the underlying mechanisms remain unclear ^22^. We also showed, as hypothesized, that adults with type 1 diabetes had smaller TBV, WMV, and volumes of cortex, nucleus accumbens, and thalamus, compared to matched controls. Earlier, at the average age of 40, with disease duration of 22 years, the same cohort showed differences only in thalamus volume, compared with healthy controls ^29^. The more pronounced differences between individuals with type 1 diabetes and controls are likely due to the longer exposure for diabetic risk factors, as type 1 diabetes has been shown to accelerate brain aging by 4–9 years ^3^. Our results are consistent with previous studies reporting differences between controls and middle-aged adults with type 1 diabetes in TBV ^3^, WMV ^3,9^, GMV ^3,13^, and thalamic ^2^ and cortical ^10^ volumes. However, cortical volume differences have not been consistently observed ^11,14^, possibly because of smaller sample sizes in earlier studies. No significant differences in hippocampal volume were found between individuals with type 1 diabetes and controls, consistent with most earlier studies ^10,36^ and aligning with the general absence of memory deficits ^8^, a characteristic of hippocampal degeneration, in adults with type 1 diabetes during midlife. One study reported smaller right hippocampal volume in older type 1 diabetes individuals with a longer disease duration, which, with methodology differences, may account for the discrepancy ^12^.

Effect sizes for volumetric differences were larger when comparing type 1 diabetes individuals with and without CMB burden than when comparing type individuals with 1 diabetes and controls. Similar finding was observed in the same cohort already six years prior, when comparing individuals with type 1 diabetes and with and without cSVD markers ^29^. One possibility is that cSVD may itself contribute to volume loss, as it is known to contribute to brain atrophy in older people ^37^. A longitudinal study in middle-aged adults with type 1 diabetes showed cortical structural changes over 3.5 years, with higher follow-up HbA_1c_ associated with greater left hemispheric surface area loss ^16^.

Other possible mechanisms may include insulin resistance, and immunological and inflammatory factors ^38^. In our study, insulin resistance was associated with the TBV, and cortex and choroid plexus volumes, consistently with earlier studies showing associations between vascular and glycemic risk factors and brain volumes ^3,11,15,16^, even though also contradictory evidence exists for GMVs ^15,39^. Microvascular complications, including retinopathy and low eGFR, were not associated with any volumetric measure, in line with several earlier reports ^3,11^. This may reflect our exclusion criteria resulting in a sample with relatively mild complications ^18^, or that microvascular complications share variance with HbA_1c_ when entered into the same model.

Contrary to our hypothesis, brain volumes were not directly associated with cognition in individuals with type 1 diabetes. This differs from earlier studies reporting associations between brain volume loss and poorer cognitive performance across multiple regions and cognitive domains ^3,9,11,12^. The differing results may reflect earlier study cohorts’ smaller sample sizes ^9^, older age ^3^, or longer diabetes duration ^3,12^ compared to our cohort.

Importantly however, we showed for the first time a negative synergistic effect of CMBs, a marker of cSVD, and brain atrophy on cognition. Compared to individuals with none or few CMBs, those with several CMBs manifested associations between TBV, WMV, nucleus accumbens, and choroid plexus volume and processing speed and executive function performance. Only one earlier study has examined the moderating role of cSVD, as indicated with WMHs, on the association between GMV and cognition, finding no effect ^12^. The finding that choroid plexus enlargement was more strongly associated with poorer cognitive performance among those with larger CMB burden is in accordance with older general population with cSVD ^21^ and individuals with type 2 diabetes ^34^.

The cross-sectional nature of the study prevents us from assessing how the volumetric changes would relate to cognitive changes over time. A further limitation is the overall sample size, which may be insufficient to detect the expectedly small effect sizes for associations between brain volumes and cognition in middle age. Specifically, the subgroup with ≥3 CMBs was relatively small, limiting the reliability of interaction analyses and warranting replication in larger cohorts, though we identified no influential data points affecting the results. Additionally, generalizability to all individuals with type 1 diabetes may be restricted, as our sample was limited to Finnish population not having prior neurovascular disease symptoms. Strengths of the study comprise the comprehensive clinical evaluation, standardized evaluation of cSVD, automated MRI segmentation of structural brain volumes, the use of standard neuropsychological tests, as well as sensitive computerized cognitive measures, correction of multiple testing, and reporting of standardized effect sizes.

To conclude, we found smaller global and regional brain volumes and larger choroid plexuses in individuals with type 1 diabetes already in middle age compared to healthy controls. In individuals with type 1 diabetes, CMB burden increased the likelihood of global and regional brain atrophy and enlargement of choroid plexus, and related cognitive deficits.

## Acknowledgements

We are thankful to research psychologists Tuuli Levänen, Emma Talvitie, Enni Hannukkala, and Heidi Heinonen for performing the neuropsychological assessments. We also thank research nurses Anu Dufva, Anna Sandelin, and Kirsi Uljala for technical assistance and Anne Komsi and radiographers at the New Children’s Hospital for work on brain MRI. We are grateful to all participants attending this study.

## Data availability

Individual-level data from the study participants are not publicly available due to consent restrictions provided by the participants at the time of data collection. Readers may propose collaboration to access the individual-level data by contacting the lead investigator.

## Author Contributions and Guarantor Statement

IK, LK, T-BC, JM, LMT, JP, DG, and HJ contributed to the study design, acquisition and interpretation of data. IK prepared first draft of the study. All authors contributed to the interpretation of the results and gave their critical comments for the manuscript. All authors agreed on the final content. HJ is the guarantor of this study and takes responsibility for the integrity of the data and the accuracy of the data analysis.

## Prior Presentation

Parts of this study were presented in abstract and poster form at the International Neuropsychology Society’s Mid-Year Meeting, Dublin, Ireland, 22–24 July 2026.

