## Supplemental data for "Brain volumes and their relationship with cerebral microbleeds and cognition in middle-aged adults with type 1 diabetes"

### SUPPLEMENTARY MATERIAL

**Supplemental Table 1.** FinnDiane Study Centers and their physicians and nurses.

| FinnDiane Study Centers | Physicians and nurses |
|---|---|
| **Anjalankoski Health Center** | S.Koivula, T.Uggeldahl |
| **Central Finland Central Hospital, Jyväskylä** | T.Forslund, A.Halonen, A.Koistinen, P.Koskiaho, M.Laukkanen, J.Saltevo, M.Tiihonen |
| **Central Hospital of Åland Islands, Mariehamn** | M.Forsen, H.Granlund, A.-C.Jonsson, B.Nyroos |
| **Central Hospital of Kanta-Häme, Hämeenlinna** | P.Kinnunen, A.Orvola, T.Salonen, A.Vähänen |
| **Central Hospital of Kymenlaakso, Kotka** | R.Paldanius, M.Riihelä, L.Ryysy |
| **Central Hospital of Länsi-Pohja, Kemi** | H.Laukkanen, P.Nyländen, A.Sademies |
| **Central Ostrobothnian Hospital District, Kokkola** | S.Anderson, B.Asplund, U.Byskata, P.Liedes, M.Kuusela, T.Virkkala |
| **City of Espoo Health Center:** | |
| **Espoonlahti** | A.Nikkola, E.Ritola |
| **Tapiola** | M.Niska, H.Saarinen |
| **Samaria** | E.Oukko-Ruponen, T.Virtanen |
| **Viherlaakso** | A.Lyytinen |
| **City of Helsinki Health Center:** | |
| **Puistola** | H.Kari, T.Simonen |
| **Suutarila** | A.Kaprio, J.Kärkkäinen, B.Rantaeskola |
| **Töölö** | P.Kääriäinen, J.Haaga, A-L.Pietiläinen |
| **City of Hyvinkää Health Center** | S.Klemetti, T.Nyandoto, E.Rontu, S.Satuli-Autere |
| **City of Vantaa Health Center:** | |
| **Korso** | R.Toivonen, H.Virtanen |
| **Länsimäki** | R.Ahonen, M.Ivaska-Suomela, A.Jauhiainen |
| **Martinlaakso** | M.Laine, T.Pellonpää, R.Puranen |
| **Myyrmäki** | A.Airas, J.Laakso, K.Rautavaara |
| **Rekola** | M.Erola, E.Jatkola |
| **Tikkurila** | R.Lönnblad, A.Malm, J.Mäkelä, E.Rautamo |
| **Heinola Health Center** | P.Hentunen, J.Lagerstam |
| **Helsinki University Hospital, Department of Medicine, Division of Nephrology** | R.Bergdal, T.Claesson, A.Dufva, N.Elonen, M.Eriksson, J.Fagerudd, M.Feodoroff, D.Gordin, P.-H.Groop, O.Heikkilä, K.Hietala, S.Hägg-Holmberg, F.Jansson Sigfrids, M.Korolainen, J.Kytö, S.Lindh, J.Nicklén, H.Paajanen, K.Pettersson-Fernholm, K.Rimpeläinen, M.Rosengård-Bärlund, M.Rönnback, L.Salovaara, A.Sandelin, M.Saraheimo, S.Satuli-Autere, R.Simonsen, P.Smidtslund, L.Thorn, H.Tikkanen, J.Tuomikangas, A.Tynjälä, K.Uljala, T.Vesisenaho, J.Wadén, A.Ylinen |
| **Herttoniemi Hospital, Helsinki** | V.Sipilä |
| **Hospital of Lounais-Häme, Forssa** | T.Kalliomäki, J.Koskelainen, R.Nikkanen, N.Savolainen, H.Sulonen, E.Valtonen |
| **Hyvinkää Hospital** | L. Norvio, A.Hämäläinen |
| **Iisalmi Hospital** | E.Toivanen |
| **Jokilaakso Hospital, Jämsä** | A.Parta, I.Pirttiniemi |

| | |
|---|---|
| **Jorvi Hospital, Helsinki University Central Hospital** | S.Aranko, S.Ervasti, R.Kauppinen-Mäkelin, A.Kuusisto, T.Leppälä, K.Nikkilä, L.Pekkonen |
| **Jyväskylä Health Center, Kyllö** | K.Nuorva, M.Tiihonen |
| **Kainuu Central Hospital, Kajaani** | S.Jokelainen, K.Kananen, M.Karjalainen, P.Kemppainen, A-M.Mankinen, A.Reponen, M.Sankari |
| **Kerava Health Center** | H.Stuckey, P.Suominen |
| **Kirkkonummi Health Center** | A.Lappalainen, M.Liimatainen, J.Santaholma |
| **Kivelä Hospital, Helsinki** | A.Aimolahti, E.Huovinen |
| **Koskela Hospital, Helsinki** | V.Ilkka, M.Lehtimäki |
| **Kotka Health Center** | E.Pälikkö-Kontinen, A.Vanhanen |
| **Kouvola Health Center** | E.Koskinen, T.Siitonen |
| **Kuopio University Hospital** | E.Huttunen, R.Ikäheimo, P.Karhapää, P.Kekäläinen, M.Laakso, T.Lakka, E.Lampainen, L.Moilanen, S. Tanskanen, L.Niskanen, U.Tuovinen, I.Vauhkonen, E.Voutilainen |
| **Kuusamo Health Center** | T.Kääriäinen, E.Isopoussu |
| **Kuusankoski Hospital** | E.Kilkki, I.Koskinen, L.Riihelä |
| **Laakso Hospital, Helsinki** | T.Meriläinen, P.Poukka, R.Savolainen, N.Uhlenius |
| **Lahti City Hospital** | A.Mäkelä, M.Tanner |
| **Lapland Central Hospital, Rovaniemi** | L.Hyvärinen, K.Lampela, S.Pöykkö, T.Rompasaari, S.Severinkangas, T.Tulokas |
| **Lappeenranta Health Center** | P. Erola, L.Härkönen, P.Linkola, T.Pekkanen, I.Pulli, E.Repo |
| **Lohja Hospital** | T.Granlund, K.Hietanen, M.Porrassalmi, M.Saari, T.Salonen, M.Tiikkainen, |
| **Länsi-Uusimaa Hospital, Tammisaari** | I.-M.Jousmaa, J.Rinne |
| **Loimaa Health Center** | A.Mäkelä, P.Eloranta |
| **Malmi Hospital, Helsinki** | H.Lanki, S.Moilanen, M.Tilly-Kiesi |
| **Mikkeli Central Hospital** | A.Gynther, R.Manninen, P.Nironen, M.Salminen, T.Vänttinen |
| **Mänttä Regional Hospital** | I.Pirttiniemi, A-M.Hänninen |
| **North Karelian Hospital, Joensuu** | U-M.Henttula, P.Kekäläinen, M.Pietarinen, A.Rissanen, M.Voutilainen |
| **Nurmijärvi Health Center** | A.Burgos, K.Urtamo |
| **Oulaskangas Hospital, Oulainen** | E.Jokelainen, P-L.Jylkkä, E.Kaarlela, J.Vuolaspuro |
| **Oulu Health Center** | L.Hiltunen, R.Häkkinen, S.Keinänen-Kiukaanniemi |
| **Oulu University Hospital** | R.Ikäheimo |
| **Päijät-Häme Central Hospital** | H.Haapamäki, A.Helanterä, S.Hämäläinen, V.Ilvesmäki, H.Miettinen |
| **Palokka Health Center** | P.Sopanen, L.Welling |
| **Pieksämäki Hospital** | V.Sevtsenko, M.Tamminen |
| **Pietarsaari Hospital** | M-L.Holmbäck, B.Isomaa, L.Sarelin |
| **Pori City Hospital** | P.Ahonen, P.Merisalo, E.Muurinen, K.Sävelä |
| **Porvoo Hospital** | M.Kallio, B.Rask, S.Rämö |
| **Raahe Hospital** | A.Holma, M.Honkala, A.Tuomivaara, R.Vainionpää |
| **Rauma Hospital** | K.Laine, K.Saarinen, T.Salminen |
| **Riihimäki Hospital** | P.Aalto, E.Immonen, L.Juurinen |
| **Salo Hospital** | A.Alanko, J.Lapinleimu, P.Rautio, M.Virtanen |
| **Satakunta Central Hospital, Pori** | M.Asola, M.Juhola, P.Kunelius, M.-L.Lahdenmäki, P.Pääkkönen, M.Rautavirta |

| | |
|---|---|
| **Savonlinna Central Hospital** | T.Pulli, P.Sallinen, M.Taskinen, E.Tolvanen, T.Tuominen, H.Valtonen, A.Vartia, S-L.Viitanen |
| **Seinäjoki Central Hospital** | O.Antila, E.Korpi-Hyövälti, T.Latvala, E.Leijala, T.Leikkari, M.Punkari N.Rantamäki, H.Vähävuori |
| **South Karelia Central Hospital, Lappeenranta** | T.Ensala, E.Hussi, R.Härkönen, U.Nyholm, J.Toivanen |
| **Tampere Health Center** | A.Vaden, P.Alarotu, E.Kujansuu, H.Kirkkopelto-Jokinen, M.Helin, S.Gummerus, L.Calonius, T.Niskanen, T.Kaitala, T.Vatanen |
| **Tampere University Hospital** | P. Hannula, I.Ala-Houhala, R.Kannisto, T.Kuningas, P.Lampinen, M.Määttä,H.Oksala, T.Oksanen, A.Putila, H.Saha, K.Salonen, H.Tauriainen, S.Tulokas |
| **Tiirismaa Health Center, Hollola** | T.Kivelä, L.Petlin, L.Savolainen |
| **Turku Health Center** | A.Artukka, I.Hämäläinen, L.Lehtinen, E.Pyysalo, H.Virtamo, M.Viinikkala, M.Vähätalo |
| **Turku University Central Hospital** | K.Breitholz, R.Eskola, K.Metsärinne, U.Pietilä, P.Saarinen, R.Tuominen, S.Äyräpää |
| **Vaajakoski Health Center** | K.Mäkinen, P.Sopanen |
| **Valkeakoski Regional Hospital** | S.Ojanen, E.Valtonen, H.Ylönen, M.Rautiainen, T.Immonen |
| **Vammala Regional Hospital** | I.Isomäki, R.Kroneld, L.Mustaniemi, M.Tapiolinna-Mäkelä |
| **Vasa Central Hospital** | S.Bergkulla, U.Hautamäki, V-A.Myllyniemi, I.Rusk |

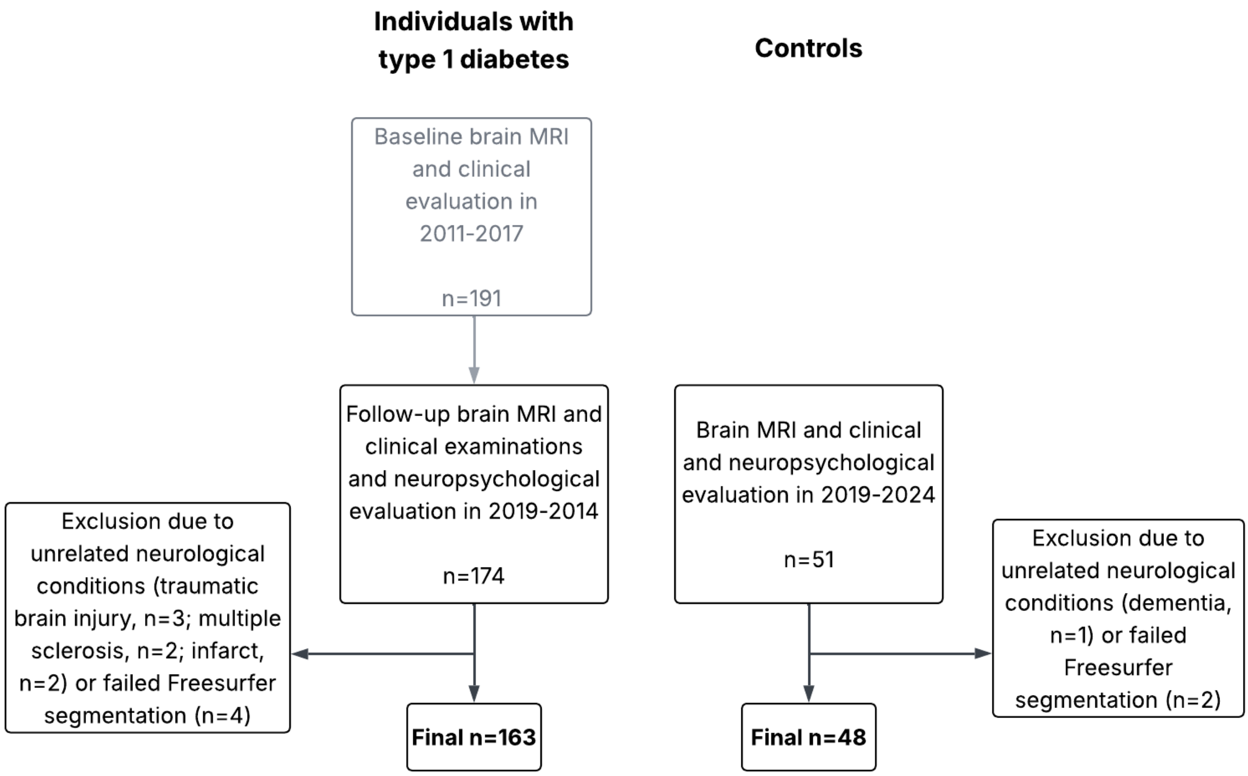


**Supplemental Figure 1.** Flowchart of the participant recruitment and data acquisition for the current study.

**Supplemental Table 2.** Neuropsychological test variables.

| | | Description of the test | Variables | Cognitive function |
|---|---|---|---|---|
| **WAIS-IV Coding** [1] | | A sequence of numbers, each paired with a corresponding hieroglyphic-like symbol. Using a key, the examinee writes the symbol corresponding to its number. | Number of correct responses within a 2-minute time limit | Processing speed |
| **Flexible Attention Test** [2,3] | | | | |
| 1 | Reaction Time | 24 light grey circles are randomly distributed on the touch screen. One circle at the time turns green. The task is to tap the green circle as quickly as possible. | Total time to complete the task (s) | Processing speed |
| 2 | Numbers and Months Backward | Numbers 1-12 and months Jan-Dec are randomly distributed in circles on the touch screen. The task is to alternately tap numbers and months in backward order in a sequence of 1-Dec-2-Nov-3-Oct, etc. | Total time to complete the task (s) divided by number of correct responses (max. 24) | Executive functions (Cognitive flexibility) |
| 3 | Visuospatial Memory Span Backward | Corsi Block-Tapping type backward span task: 24 light grey circles are distributed on the screen, one at a time turning red. The task is to tap the same circles, but in reverse order. The length of the sequence to be recalled gradually increases until the subject makes two consecutive errors in the same span length. | Maximum backward span score | Executive functions (Visuospatial working memory) |
| **Stroop Color-Naming** [4,5] | | 100 colors (written as XXXX) are printed in either red, green, blue, or yellow. The task is to name the colors as fast as possible. | Total time to complete the task (s) | Processing speed |
| **Stroop Color-Incongruent** [4,5] | | 100 colored words are written so that no word for a color matches the ink color (e.g., the word "blue" printed in red ink). The task is to name the ink color of the word as fast and precise as possible, while inhibiting the word meaning. | Total time to complete the task (s) divided by number of correct responses (max. 100) | Executive functions (Inhibition) |

**Supplemental Table 3.** Associations between brain volumes and demographic and clinical factors in individuals with type 1 diabetes.

| | Total brain volume | White matter | Cortex | Thalamus | Hippo-campus | Nucleus Accumbens | Choroid Plexus |
|---|---|---|---|---|---|---|---|
| **Age** | **-0.44 (<0.001)** | **-0.25 (0.005)** | **-0.36 (<0.001)** | **-0.37 (<0.001)** | **-0.29 (<0.001)** | **-0.26 (0.002)** | 0.15 (0.079) |
| **Female Sex** | 0.19 (0.171) | -0.14 (0.394) | **0.28 (0.036)** | 0.22 (0.152) | **0.66 (<0.001)** | 0.02 (0.899) | 0.29 (0.078) |
| **eGDR** | **0.19 (0.019)** | 0.03 (0.739) | **0.30 (<0.001)** | 0.07 (0.449) | -0.05 (0.600) | 0.16 (0.081) | **-0.22 (0.021)** |

Multivariable linear regression models with normalized brain MRI volumes as dependent variables. Independent variables included age, sex, and estimated glucose disposal rate (eGDR). Values are standardized beta coefficients (p-values). For sex, β reflects female vs. male.

**Supplemental Table 4.** Associations between brain volumes and demographic and clinical factors in individuals with type 1 diabetes.

| | Total brain volume | White matter | Cortex | Thalamus | Hippo-campus | Nucleus Accumbens | Choroid Plexus |
|---|---|---|---|---|---|---|---|
| **Age** | **-0.47 (<0.001)** | **-0.26 (0.007)** | **-0.40 (<0.001)** | **-0.33 (<0.001)** | **-0.28 (0.002)** | **-0.28 (0.002)** | 0.12 (0.195) |
| **Female Sex** | 0.17 (0.235) | -0.14 (0.426) | 0.25 (0.068) | 0.27 (0.088) | **0.66 (<0.001)** | 0.03 (0.877) | 0.26 (0.135) |
| **eGDR** | **0.19 (0.025)** | 0.01 (0.895) | **0.30 (<0.001)** | 0.04 (0.667) | -0.04 (0.669) | 0.13 (0.153) | **-0.20 (0.041)** |
| **eGFR** | -0.09 (0.225) | -0.04 (0.688) | -0.10 (0.155) | 0.09 (0.270) | 0.02 (0.781) | -0.06 (0.449) | -0.08 (0.395) |
| **Diabetic retinopathy** | 0.17 (0.252) | 0.22 (0.212) | 0.12 (0.380) | 0.12 (0.453) | -0.10 (0.545) | **0.33 (0.048)** | -0.07 (0.677) |

Multivariable linear regression models with normalized brain MRI volumes as dependent variables. Independent variables included brain age, sex, estimated glucose disposal rate (eGDR), estimated glomerular filtration rate (eGFR), and photocoagulation status. Values are standardized beta coefficients (p-values). For sex, β reflects female vs. male.

**Supplemental Table 5.** Associations between brain volumes and cognition in individuals with type 1 diabetes.

| | Processing speed | | | Executive functions | | |
|---|---|---|---|---|---|---|
| | **Coding** | **FAT-RT** | **Stroop-II** | **Stroop-III** | **FAT-NMB** | **FAT-MB** |
| **TBV** | 0.08 (0.387) | 0.04 (0.671) | -0.02 (0.865) | 0.04 (0.631) | 0.05 (0.609) | -0.01 (0.951) |
| **Age** | **-0.36 (<0.001)** | **-0.49 (<0.001)** | **-0.22 (0.018)** | **-0.27 (0.004)** | **-0.37 (<0.001)** | -0.15 (0.128) |
| **Female sex** | 0.17 (0.256) | -0.08 (0.575) | 0.09 (0.588) | 0.09 (0.567) | -0.23 (0.140) | **-0.51 (0.003)** |
| **White matter** | 0.11 (0.132) | 0.01 (0.930) | -0.01 (0.895) | 0.05 (0.527) | 0.08 (0.318) | -0.07 (0.416) |
| **Age** | **-0.38 (<0.001)** | **-0.50 (<0.001)** | **0.21 (0.011)** | **-0.28 (<0.001)** | **-0.38 (<0.001)** | -0.17 (0.053) |
| **Female sex** | 0.21 (0.148) | -0.07 (0.631) | -0.08 (0.598) | 0.11 (0.475) | -0.20 (0.178) | **-0.51 (0.002)** |
| **Cortex** | 0.03 (0.730) | 0.04 (0.616) | 0.03 (0.787) | 0.01 (0.955) | 0.02 (0.784) | 0.13 (0.190) |
| **Age** | **-0.39 (<0.001)** | **-0.48 (<0.001)** | **-0.20 (0.030)** | **-0.29 (0.002)** | **-0.39 (<0.001)** | -0.08 (0.379) |
| **Female sex** | 0.18 (0.235) | -0.09 (0.541) | 0.07 (0.675) | 0.10 (0.530) | -0.22 (0.155) | **-0.57 (<0.001)** |
| **Thalamus** | 0.07 (0.385) | 0.14 (0.074) | -0.03 (0.759) | -0.06 (0.499) | 0.00 (0.998) | 0.11 (0.246) |
| **Age** | **-0.38 (<0.001)** | **-0.45 (<0.001)** | **-0.22 (0.010)** | **-0.31 (<0.001)** | **-0.40 (<0.001)** | -0.10 (0.286) |
| **Female sex** | 0.18 (0.227) | -0.11 (0.448) | 0.09 (0.577) | 0.12 (0.445) | -0.21 (0.166) | **-0.54 (0.001)** |
| **Hippocampus** | -0.03 (0.733) | -0.06 (0.456) | -0.11 (0.213) | -0.06 (0.501) | -0.01 (0.917) | 0.09 (0.318) |
| **Age** | **-0.42 (<0.001)** | **-0.52 (<0.001)** | **-0.25 (0.003)** | **-0.31 (<0.001)** | **-0.40 (<0.001)** | -0.12 (0.174) |
| **Female sex** | 0.21 (0.167) | -0.03 (0.835) | 0.15 (0.373) | 0.14 (0.394) | -0.20 (0.200) | **-0.56 (0.001)** |
| **Nucleus accumbens** | 0.02 (0.803) | 0.11 (0.154) | 0.02 (0.775) | 0.15 (0.068) | 0.00 (0.964) | 0.07 (0.420) |
| **Age** | **-0.40 (<0.001)** | **-0.47 (<0.001)** | **-0.21 (0.015)** | **-0.24 (0.003)** | **-0.40 (<0.001)** | -0.12 (0.153) |
| **Female sex** | 0.19 (0.186) | -0.08 (0.545) | 0.08 (0.619) | 0.08 (0.584) | -0.21 (0.164) | **-0.52 (0.002)** |
| **Choroid plexus** | 0.00 (0.976) | -0.01 (0.842) | -0.06 (0.429) | -0.08 (0.336) | 0.00 (0.986) | -0.03 (0.727) |
| **Age** | **-0.41 (<0.001)** | **-0.50 (<0.001)** | **-0.03 (0.017)** | **-0.27 (<0.001)** | **-0.40 (<0.001)** | -0.14 (0.104) |
| **Female sex** | 0.20 (0.182) | -0.07 (0.638) | 0.09 (0.559) | 0.12 (0.451) | -0.21 (0.162) | **-0.50 (0.003)** |

Linear regression models with cognitive test scores as dependent variables. Independent variables included brain MRI volume marker, age, and sex. Values are standardized beta coefficients (p-values). For sex, β reflects female vs. male. FAT, Flexible Attention Test; FAT-MB, FAT Visuospatial Memory Span Backward; FAT-NMB, FAT Numbers and Months Backward; FAT-RT, FAT Reaction Time; Stroop-II, Stroop Color-Naming; Stroop-III, Stroop Color-Incongruent; TBV, total brain volume. Z-scores for FAT-RT, FAT-NMB, Stroop-II, and Stroop-III are reversed so that higher scores consistently indicate better performance across all cognitive tests.

**Supplemental Table 6.** Interactions of brain volumes and cerebral microbleeds (CMBs) on cognition in individuals with type 1 diabetes.

| | **Processing speed** | | | **Executive functions** | | |
|---|---|---|---|---|---|---|
| | **Coding** | **FAT-RT** | **Stroop-II** | **Stroop-III** | **FAT-NMB** | **FAT-MB** |
| TBV × CMBs | 0.28 (0.215) | **0.54 (0.013)**† | 0.23 (0.351) | **0.48 (0.045)** | **0.75 (0.001)**† | 0.14 (0.558) |
| White matter × CMBs | 0.38 (0.088) | **0.65 (0.002)**† | 0.43 (0.076) | **0.47 (0.048)** | **0.68 (0.003)**† | 0.36 (0.164) |
| Cortex × CMBs | -0.06 (0.815) | 0.10 (0.694) | -0.33 (0.265) | -0.06 (0.848) | **0.68 (0.015)** | 0.07 (0.815) |
| Thalamus × CMBs | 0.38 (0.110) | 0.36 (0.114) | -0.05 (0.856) | 0.18 (0.478) | 0.05 (0.840) | 0.16 (0.516) |
| Hippocampus × CMBs | 0.06 (0.760) | 0.29 (0.135) | -0.03 (0.889) | 0.19 (0.367) | 0.31 (0.132) | -0.06 (0.786) |
| Nucleus accumbens × CMBs | 0.35 (0.082) | 0.22 (0.262) | 0.41 (0.060) | **0.56 (0.008)**† | 0.24 (0.257) | -0.10 (0.646) |
| Choroid plexus × CMBs | **-0.46 (0.027)** | **-0.61 (0.002)**† | -0.11 (0.628) | -0.11 (0.621) | **-0.51 (0.016)**† | -0.17 (0.462) |

Interaction models with cognitive test scores as dependent variables. Interaction term included CMBs (0-2 or ≥3) × brain volume marker and the model was corrected for age and sex. Values are standardized beta coefficients (p-values) for the interaction terms. FAT, Flexible Attention Test; FAT-MB, FAT Visuospatial Memory Span Backward; FAT-NMB, FAT Numbers and Months Backward; FAT-RT, FAT Reaction Time; Stroop-II, Stroop Color-Naming; Stroop-III, Stroop Color-Incongruent; TBV, total brain volume. Z-scores for FAT-RT, FAT-NMB, Stroop-II, and Stroop-III are reversed so that higher scores consistently indicate better performance across all cognitive tests. Significant interactions after false discovery rate correction are marked with †.
